# Telemedicine-Based Buprenorphine Initiation and Maintenance in Rural Jails: A Retrospective Observational Study

**DOI:** 10.64898/2026.01.29.26345153

**Authors:** Annabelle M. Belcher, Allison O’Rourke, Hannah C. Smith, Heather Fitzsimons, Kristianny Ruelas-Vargas, Christopher J. Welsh, Brendan Saloner, Eric Weintraub

**Affiliations:** Division of Addiction Research and Treatment, Department of Psychiatry, University of Maryland School of Medicine, Baltimore, MD 21201, USA; Department of Behavioral and Social Sciences, Brown University School of Public Health, Providence, RI 02903

## Abstract

**BACKGROUND:** This study evaluates the reach, scalability, and implementation of a large-scale, multi-site tele-buprenorphine program designed to treat opioid use disorder (OUD) within rural carceral settings. Given that individuals transition frequently between jails and the community, these facilities represent a critical window for OUD intervention, yet they often face significant provider shortages and logistical barriers. We conducted a retrospective chart review of 842 unique patients (1,321 treatment episodes) enrolled in the University of Maryland’s tele-buprenorphine program across six rural county jails between June 2020 and May 2025. Data extracted from jail records and electronic health records were used to analyze patient demographics, prescribing patterns, and program retention.

**RESULTS:** The patient population was primarily male (71.1%) and White (75.7%), with a mean age of 35.4 years. Participants reported high-severity OUD, with an average of 12.6 years of opioid use. Reflecting broad admission criteria, 55.2% of participants were new treatment initiates not receiving MOUD prior to booking. Patients spent a mean of 35.6 days incarcerated before initiation and were retained in the program for an average of 66 days. Buprenorphine doses were titrated from a mean initiation dose of 8.8 mg to 16.2 mg at discharge. The program demonstrated a 99.5% adherence rate among retained patients. Only 3% of the total sample were discharged for medication diversion or hoarding.

**CONCLUSIONS:** Telemedicine is a highly feasible and scalable model for delivering evidence-based MOUD in rural jails. By utilizing a “liberal admission policy” that prioritizes both treatment initiation and maintenance, programs can successfully reach high-risk individuals who lack access to community-based care. These findings suggest that tele-buprenorphine can effectively bridge the treatment gap in underserved jurisdictions, potentially reducing the risk of overdose during the high-risk post-release period.

## BACKGROUND

For over a decade, the United States has been grappling with a persistent opioid epidemic (*Products - Vital Statistics Rapid Release - Provisional Drug Overdose Data*, 2025)—a public health crisis that has had a disproportionate impact on people who are incarcerated. Estimates suggest that up to 36% of people with opioid use disorder (OUD) cycle through the correctional system each year (Boutwell et al., 2007), and that 15% of people who are incarcerated have an OUD (Lenz et al., 2025). Incarceration also poses a high risk of overdose: formerly incarcerated individuals are significantly more likely to die of a drug overdose in the first days to weeks following release from jail or prison (Merrall et al., 2010). Post-release overdose has historically been a major problem in Maryland, a state that has been hard hit by the opioid epidemic (Maryland Department of Health and Mental Hygiene, 2014). The reasons for this heightened risk are many and include multiple social determinants of health (Berk et al., 2025); but it is widely known that many people undergo forced abstinence in jails and prisons, leading to a higher risk of overdose upon release (ASAM, 2020; Rich et al., 2015).

The provision of medications for opioid use disorder (MOUD), such as buprenorphine, to incarcerated individuals is associated with a host of improved public health outcomes (Macmadu et al., 2020; National Academies of Sciences, Engineering, and Medicine, 2019), including a decrease in post-release overdose deaths (Evans, Wilson, et al., 2022; Friedmann et al., 2025; Green et al., 2018; Lim et al., 2022) and increased community treatment engagement (Friedmann et al., 2025; Gordon et al., 2014; Magura et al., 2009). Jails serve as short-term detention stays for individuals awaiting trial, an ideal window within which to provide MOUD treatment. Additionally, with a high population turnover (Zeng, 2022), jails serve as a direct pipeline back to the local community. For these reasons, jails present a unique opportunity for MOUD provision. But implementation is frequently hindered by logistical challenges, including medication dosing expertise, staffing resources, and space constraints (Flanagan Balawajder et al., 2024). Addressing these barriers represents a critical opportunity to expand treatment access, particularly for rural, geographically isolated and medically underserved areas.

Telemedicine offers a scalable approach to mitigate persistent healthcare and treatment disparities in rural areas (Rubin, 2019) and may have a unique role for OUD treatment provision in correctional facilities. COVID-19 accelerated the implementation of telemedicine and demonstrated that it could be an effective and safe way to expand MOUD access (Donelan et al., 2021; Duncan et al., 2021; Zaller & Brinkley-Rubinstein, 2020), but documented evidence of its application for delivering MOUD as standard practice within carceral settings is scarce. The problem of access is particularly acute in rural communities (Haffajee et al., 2019; Hedegaard et al., 2019; Lister et al., 2020; Morenz et al., 2025), where resource and healthcare deficits exacerbate the inherent challenges of MOUD access in local jails.

The University of Maryland School of Medicine Division of Addiction Research and Treatment (UM DART) has established expertise providing telemedicine to a variety of community clinical settings (Weintraub et al., 2018; Weintraub, Greenblatt, et al., 2021; Weintraub, Seneviratne, et al., 2021). In 2019, the state of Maryland passed House Bill (HB) 116 mandating that all local correctional facilities make at least one formulation of each FDA– approved full opioid agonist, partial opioid agonist, and long–acting opioid antagonist used for the treatment of opioid use disorders available to any incarcerated individual in need (Maryland Correctional Services Code Ann. §9-603, 2020). Pushed by the momentum of the new Maryland law, UM DART applied for and was awarded private foundation funding to launch a new clinical program to provide treatment for OUD with buprenorphine via telemedicine (tele-buprenorphine) to patients incarcerated in rural county detention centers (FORE Foundation, 2020). The initial single-site program demonstrated successful implementation which enabled patients to begin and engage with tele-buprenorphine treatment before the critical, high-risk window following release (Belcher et al., 2021). Further, a recent single-site evaluation of the program at the highest-volume jail confirms a high rate of post-release treatment engagement, with 78% of patients remaining in buprenorphine treatment within two weeks of discharge (Belcher et al., 2025). However, real-world data are needed to understand the broader program’s potential reach, scale, and implementation. Here we report patient characteristics, medication prescription, and program retention of all first encounters of patients in the UM tele-buprenorphine program seen during the program’s first five years of operation.

## METHODS

### Study Design

We conducted a retrospective chart review of first admissions of incarcerated individuals who were enrolled in one of six locations throughout the state of Maryland for the tele-buprenorphine program and released between June 1, 2020 and May 31, 2025. All facilities were located in rural counties as designated by the Maryland State Office of Rural Health with average daily census ranging from approximately 40 to 160. The majority of the populations in both the counties and jails identify as White (Table 1). County overdose mortality rates ranged from 12.0 to 73.1 overdose deaths annually per 100,000 population. The study was approved as a consent-waivered chart review by the University of Maryland’s (UM) Human Research Protection Office (UMB IRB protocol No. HP - 00100535). This study is reported using Strengthening the Reporting of Observational Studies in Epidemiology (STROBE) guidelines.

**Table 1.** Jail and Detention Center (Study Site) Characteristics.

| **County Detention Center** | **Facility Average Daily Census** | **County Zip code; RUCA Code** | **County % White\*** | **County Overdose Mortality Rate (per 100,000)\*** | **Tele-bupe Program Initiation Date** | **Tele-bupe Total Number of Unique Patients** | **Tele-bupe % White** |
| --- | --- | --- | --- | --- | --- | --- | --- |
| **Allegany** | 162 | Rural | 88% | 66.1 | 4/23/21 | 446 | 80% |
| **Caroline** | 57 | Rural | 81% | 42.1 | 2/20/23 | 80 | 75% |
| **Kent** | 49 | Rural | 81% | 20.8 | 1/19/22 | 49 | 65% |
| **Queen Anne's** | 60 | Rural | 90% | 12.0 | 5/10/23 | 81 | 75% |
| **Somerset** | 40 | Urban | 55% | 73.1 | 12/15/22 | 79 | 59% |
| **Talbot** | 60-80 | Rural | 83% | 18.7 | 6/15/20 | 51 | 73% |

Characteristics of jails and detention center sites that receive tele-buprenorphine services through the University of Maryland program. Carceral facilities were matched by zip code to 2020 US Department of Agriculture Rural-Urban Commuting Area (RUCA) codes to delineate rurality. RUCA codes classify areas by zip codes on a scale from 1, most metropolitan, to 10, rural, based on population and proximity to core metropolitan areas. For this table urban and rural categories were created as: Urban = RUCA 1-3 and Rural=RUCA4-10; \*Connotes data obtained from the 2020 Census.

### Setting and Participants

Implementation of this program has been described previously (Belcher et al., 2021; Spaderna et al., 2025), but briefly: tele-buprenorphine was established in six rural county jails in Maryland. Upon incarceration, individuals are given a brief screen for OUD by jail medical staff, the designated on-site contractual nurse provider, or the designated Medications for Addiction Treatment (MAT) nurse, and if positive, provided with comfort medications prior to evaluation by a UM provider via HIPAA-compliant video conferencing. Following the provider encounter, patients are prescribed daily buprenorphine at a therapeutically appropriate dose (usually between 16 mg to 24 mg), supplied as 2 mg or 8 mg tablets (or a combination thereof). Daily dosing is provided by the designated nurse. Follow-up encounters are scheduled either bi-weekly or monthly, as clinically indicated, with services provided at release to support care continuity in the community through a referral and (typically) a one-week bridge prescription.

### Data Sources and Data Collection

At enrollment, patients signed a release of information to allow the UM treatment team to access their jail records. Data were collected from jail intake and discharge records and the electronic health record (EHR) of treated patients. Jail intake records included patient characteristics at admission (intake into the jail) including demographics and substance use history, in addition to dates related to booking and discharge.

EHR records included medical care items related to pre-incarceration MOUD treatment, and program measures related to buprenorphine treatment while incarcerated. Using a standardized extraction tool, trained UM research staff abstracted data from these two sources following patient jail discharge. De-identified data were recorded in REDCap, a HIPAA compliant data collection and management software (Harris et al., 2009).

### Outcome Measures

Measures in our analyses included patient characteristics at admissions, pre-treatment clinical characteristics, tele-buprenorphine prescribing during incarceration, and tele-buprenorphine program measures. Reported patient characteristics include sex (male, female, other), age at correctional intake, race (Caucasian/White, African American/Black, Native American/Alaska Native, Asian/Asian American, Unsure, Other), Hispanic/Latino ethnicity (yes/no), relationship status (married or significant other, divorced, single, widowed), and total days of incarceration. Patient self-reported their race as a check-all-that-apply measure. Due to small group sizes, race is reported for three categories: White, Black, and Other (any endorsement of race as Native America/Alaska Native, Asian/Asian American, Unsure or other). Marital status was condensed to two groups, married or significant other versus divorced, single, or widowed. Total days of incarceration was calculated during extraction as the difference, in days, between date of correctional intake and release. Pre-treatment clinical characteristics included self-reported years of opioid use at time of admission, route of most recent opioid use (insufflation, intravenous, smoke, oral), buprenorphine or methadone prescription prior to incarceration (yes/no), and number of days incarcerated prior to tele-buprenorphine initiation. Route of opioid use is reported as dichotomous for injection drug use. Days prior to tele-buprenorphine initiation was calculated as the difference between correctional intake and date of first buprenorphine dose. Tele-buprenorphine prescribing during incarceration measures include measures of mean buprenorphine dose in mg at initiation, treatment week 1 and last dose while incarcerated. Tele-buprenorphine program measures include program retention and program adherence.

Program retention is defined as patients who were documented as having been initiated on buprenorphine through the tele-buprenorphine program and maintained until their release from custody. Patients that were discharged, either for non-adherence to jail program rules (e.g., diversion, hoarding), self-discharge or other were coded as discharged, and the reason for discharge was documented. Program adherence was defined as having no missed or refused buprenorphine doses during program enrollment.

### Sample Size and Statistical Analysis

Our total analytical sample consisted of 842 patients who received buprenorphine through the tele-buprenorphine program between June 2020 and May 2025. These patients included 786 patients who were initiated on buprenorphine and retained in care and 56 patients who were initiated on buprenorphine and discharged during incarceration. Summary statistics, counts and percentage for categorical measures and mean and standard deviation for continuous measures were calculated for all outcomes by each subgroup. Bivariate comparisons using chi-square and t-tests were conducted to compare retained (n=786) and discharged patients (n=56). Further, within the sample of 786 patients who were retained in care, we made comparisons between those who reported recent MOUD community treatment (n=350) or not (n=436) enrolled in community-based MOUD agonist treatment at the time of jail booking. SAS 9.4 was used for all analyses.

## RESULTS

Table 2 presents all patient population characteristics. Patients in the tele-buprenorphine program were primarily male (72.2%) with a mean age of 35.3 years old. Patients self-identified as White (75.2%) followed by Black (23.6%), and less than 1% indicated being Hispanic or Latinx. At time of admission to the program, 75.2% identified as being divorced, single or widowed. Patients were incarcerated for an average of 108.8 days. Prior to initiating buprenorphine treatment, patients reported having used opioids for 12.5 years and almost 40% reported injection opioid use as their most recent route of administration. Most patients were not on a current treatment plan at booking (i.e., were new initiates in jail), with 39.7% reporting buprenorphine and an additional 4.6% reporting a methadone prescription prior to incarceration. Patients spent an average of 36.0 days incarcerated prior to initiation in the tele-buprenorphine program.

**Table 2.** Population Characteristics of First Admission Visits of Tele-Buprenorphine Patients (N=842).

|  | All First Admissions<br>N=842 | Retained in Care<br>N=786 | Discharged from Care<br>N=56 |
| --- | --- | --- | --- |
|  | N (%) | N (%) | N (%) |
| Sex |  |  |  |
| Female | 233 (27.7) | 226 (28.8) | 7 (12.5)* |
| Male | 608 (72.2) | 559 (71.1) | 49 (87.5) |
| Other | 1 (0.1) |  |  |
| Age at correctional intake [Mean (SD)] | 35.3 (8.4) | 35.4 (8.3) | 33.6 (9.4) |
| Race (Check all that apply) |  |  |  |
| Caucasian/White | 633 (75.2) | 595 (75.7) | 38 (67.9) |
| African American/Black | 199 (23.6) | 181 (23.0) | 18 (32.1) |
| Other | 12 (1.4) |  |  |
| Hispanic/Latino | 6 (0.7) |  |  |
| Relationship Status |  |  |  |
| Married or significant other | 187 (22.2) | 177 (23.1) | 10 (18.2) |
| Divorced, Single or Widowed | 633 (75.2) | 588 (76.9) | 45 (81.8) |
| Total Length of Stay (Days of Incarceration; Mean [SD]) | 108.8 (98.9) | 103.9 (95.1) | 178 (124.1)* |
| **Pre-Treatment Clinical Characteristics** |  |  |  |
| Years of Opioid Use [Mean (SD)] | 12.5 (8.2) | 12.6 (8.2) | 10.5 (8.8) |
| Most recent route of opioid use by injection | 327 (38.8) | 313 (39.8) | 14 (25.0)* |
| Buprenorphine prescription prior to incarceration | 334 (39.7) | 316 (40.4) | 18 (32.1) |
| Methadone patient prior to | 39 (4.6) | 34 (4.4) | 5 (8.9) |
| incarceration |  |  |  |
| Length of stay prior to tele-buprenorphine initiation [Days, Mean (SD)] | 36.0 (60.3) | 35.6 (60.9) | 40.6 (50.9) |
| **Tele-Buprenorphine Prescribing During Incarceration** |  |  |  |
| Buprenorphine Initiation Dose in mg [Mean (SD)] | 8.8 (6.0) | 8.8 (6.0) | 8.3 (6.3) |
| Week 1 Buprenorphine Dose in mg [Mean (SD)] | 12.8 (5.8) | 12.9 (5.9) | 11.8 (5.2) |
| Last Buprenorphine Dose in Detention Center in mg [Mean (SD)] | 16.2 (6.4) | 16.2 (6.3) | 16.9 (8.2) |
| **Tele-Buprenorphine Program Measures** |  |  |  |
| Program Retention (Days, [Mean (SD)] | 65.3 (68.0) | 65.9 (65.6) | 56.4 (59.0) |
| Program Adherence (N, [% of total sample]) | 835 (99.2) | 781 (99.5) | 54 (96.4) * |

Population characteristics for first admissions of 842 incarcerated patients treated in the University of Maryland tele-buprenorphine program from June 2020 – May 2025. \* Bivariate analyses (chi-square or t-test): Comparing Retained in Care to those Discharged from care during incarceration, Comparing those with a MOUD prescription prior to program entry versus through without prior MOUD; Significant difference ( $p < .05$ ).

During the course of tele-buprenorphine program enrollment, patient buprenorphine doses increased on average from 8.8 mg at initiation to 16.2 mg at last dose prior to discharge from incarceration. Patients were retained in the program, on average, nearly 66 days with 99.2% tele-buprenorphine program adherence. Compared with incarceration length, patients received buprenorphine 60.0% of the days they were incarcerated.

Patients discharged from care (n = 56) were removed for diversion or hoarding medications (50%), self-discharged (26.8% e.g., didn’t like how medication made them feel or had side effects), or discharged for other reasons (23.2%; e.g., repeated behavioral concerns with staff). Compared to patients who were retained in care, discharged patients were significantly more likely to be male (87.5%, p=.0085) and less likely to report injection as their most recent route for opioid use (14%, p=.0279). While the number of incarceration days prior to buprenorphine program enrollment were similar (40.6 days), length of incarceration was significantly longer for those discharged from care, 65.9 vs. 178.0 days, respectively (p<.0001).

Nearly half (44.5%) of retained patients reported a recent MOUD prescription in the days prior to incarceration (Table 3). Significant differences were found between patients with MOUD prescriptions at carceral admission and those without. Demographically, patients with MOUD prescriptions were significantly older (36.6 vs. 34.5 years old, p=.0005) and female (32.6% vs. 25.7%, p=.0319). Differences were also identified by race with a larger percentage of patients with prior MOUD prescriptions identifying as White (81.1% vs. 71.5%, p=.0014) and a smaller percentage identifying as Black (18.0% vs. 27.1%, p=.0027) compared with patients without previous MOUD prescriptions. They also experienced significantly shorter lengths of incarceration compared with patients without prior MOUD prescriptions (93.2 days vs. 112.6 days, p=.0046). Differences were also identified in time prior to program buprenorphine initiation and dosing. Patients with prior MOUD prescriptions had significantly fewer days incarcerated prior to buprenorphine initiation (21.3 days vs. 47.3 days, p=<.0001) and experienced higher buprenorphine doses. At initiation, patients with prior MOUD averaged a dose of 11.9 mg compared with non-MOUD patients at 6.3 mg (p<.0001). These higher doses persisted throughout treatment as measured at week 1 (15.1 mg vs. 11.1 mg, p<.0001) and last dose prior to discharge (17.8 mg vs, 14.9 mg, p<.0001).

**Table 3.** Retention in Care Among First Admissions (n=786) by Pre-Incarceration MOUD Status, 2020–2025.

|  | **Prior MOUD Prescription** | **No prior MOUD Prescription** |
| --- | --- | --- |
|  | **N=350** | **N=436** |
|  | **N (%)** |  |
| Sex |  |  |
| Female | 114 (32.6) | 112 (25.7)* |
| Male | 235 (67.3) | 324 (74.3) |
| Age at correctional intake [Mean (SD)] | 36.6 (7.8) | 34.5 (8.6) * |
| Race (Check all that apply) |  |  |
| Caucasian/White | 284 (81.1) | 311 (71.5) |
| African American/Black | 63 (18.0) | 118 (27.1) |
| Relationship Status |  |  |
| Married or significant other | 84 (24.5) | 93 (22.0) |
| Divorced, Single or Widowed | 259 (75.5) | 329 (78.0) |
| Total Length of Stay (Days of Incarceration; Mean [SD]) | 93.2 (90.3) | 112.6 (98.1) * |
| **Pre-Treatment Clinical Characteristics** |  |  |
| Years of Opioid Use [Mean (SD)] | 13.2 (8.3) | 12.2 (8.1) |
| Most recent route of opioid use by injection | 142 (45.4) | 171 (54.6) |
| Length of stay prior to tele-buprenorphine initiation [Mean (SD)] | 21.3 (41.9) | 47.3 (70.7) * |
| **Tele-Buprenorphine Prescribing During Incarceration** |  |  |
| Buprenorphine Initiation Dose in mg [Mean (SD)] | 11.9 (6.3) | 6.3 (4.4) * |
| Week 1 Buprenorphine Dose in mg [Mean (SD)] | 15.1 (5.4) | 11.1 (5.7) * |
| Last Buprenorphine Dose in Detention Center in mg [Mean (SD)] | 17.8 (6.0) | 14.9 (6.1) * |
| **Tele-Buprenorphine Program Measures** |  |  |
| Program Retention (Days, [Mean (SD)]) | 70.0 (74.5) | 62.6 (63.2) |

## DISCUSSION

Extensive research confirms the advantages of providing MOUD in correctional facilities, including in jails (Berk et al., 2025; Degenhardt et al., 2014; Friedmann et al., 2025); but access to this life-saving treatment in rural settings is challenged by healthcare and provider shortages. Telemedicine provides a viable pipeline for MOUD treatment and could be a particularly valuable tool in rural carceral environments. But reports of telemedicine utilization in this context are limited. Here we report the large-scale multi-site feasibility and scalability of tele-buprenorphine for treatment of incarcerated individuals. Our program successfully provided 1,321 treatment episodes to 842 unique patients across six rural Maryland county jails over five years, serving as the primary buprenorphine provider in these jails. Notably, these jails are located in rural counties in a state that, relative to the rest of the nation, has consistently reported a high overdose death rate since the program’s initiation in 2020 (approximately 40 drug overdose deaths per 100,000 total population) (CDC, 2025). The jurisdictions within our program’s reach report overdose mortality rates of up to 73.1 per 100,000 residents, further justifying the need for robust overdose prevention strategies in these locales.

Our incarcerated patient population demographic characteristics largely mirrored that of their local jurisdictions, and all were predominantly white. Patients were also predominantly male, with an average age of 35.4 years, and represented the most severe form of opioid use disorder, indicated by the high number of years of opioid use and high rate of intravenous injection use. As individuals with criminolegal involvement (short-term stays in a local rural jail with an average LOS of approximately 3.5 months) who are diagnosed with moderate-to-severe opioid use disorder, this population has been identified as being at very high risk for opioid overdose upon release into the community (Saloner et al., 2020).

Jails represent a crucial community pipeline and an ideal window for initiating MOUD treatment in a safe, controlled environment. Our team recently reported a high rate of two-week post-release community treatment engagement for patients enrolled in our jail-based tele-buprenorphine program (Belcher et al., 2025); thus, treatment initiation during incarceration is a major goal of our clinical enterprise. The UM tele-buprenorphine program was implemented from June 2020 (the first site) to May 2023 using a phased rollout strategy at each of the jail sites. As part of our programmatic implementation procedures, the team initially opened treatment exclusively to pregnant patients and to individuals requiring maintenance of prior established community treatment. Following an iterative protocol refinement period, and within three months of individual program site implementation, eligibility criteria were expanded to include all screened and referred individuals, irrespective of prior treatment history or induction status. Consistent with this open enrollment policy, the study population primarily consisted of new treatment initiates; notably, 55.2% of participants were not receiving buprenorphine or methadone prior to incarceration. This contrasts with previous observational reports of jail-based MOUD programs whose primary function is to ensure continuity of care, with the majority of services focused on maintaining treatment protocols initiated in the community (Carroll, 2025; Friedmann et al., 2025). This liberal admission policy allows the clinical team to provide services to anyone for whom treatment is determined to be appropriate, consistent with the state law (Maryland Correctional Services Code Ann. §9-603, 2020).

The UM clinical treatment team providers possessed several decades of experience providing in-person and virtual buprenorphine in community urban and rural settings with a focus on providing patient-centered care—experience which carried through to the prescribing practices that are currently employed in the rural jail tele-buprenorphine settings. The average initiation doses in this study ranged from 8 mg to 12 mg, with titration over a series of days or weeks of up to 16 mg to 24 mg. With a justified public health rationale of elucidating access issues, most reports of MOUD treatment in carceral settings focus on *whether* buprenorphine is provided in jail and prison settings at all (Flanagan Balawajder et al., 2024; Krawczyk et al., 2017). But far fewer investigations have explored the medication prescribing practices within these carceral settings, and only a handful report the exact buprenorphine medication doses (in milligrams) that are prescribed (Moore et al., 2019). In one of the clearest peer-reviewed, published assessments of carceral setting MOUD prescribing practices, a retrospective chart review of the New Jersey Department of Corrections MOUD program found that 90-day average and median doses of buprenorphine were 8.4 mg and 8 mg, respectively, with a range of 2 mg to 12 mg (Tamburello et al., 2022). Generally, buprenorphine administration in carceral settings occurs at a lower dose than that which would be provided in community settings (Grande et al., 2023), especially in light of the fentanyl crisis (Chambers et al., 2023; D’Agata Mount et al., 2024). There were several limitations to the team’s ability to provide optimal treatment focused on patient therapeutic benefit, however, including the inability to provide split-dosing regimens and the quick-judgment release of some patients who may not yet had arrived at a therapeutic buprenorphine dose (Spaderna et al., 2025). These deviations from community practice underscore the challenging nature of providing evidence-based treatment in jail settings.

This observational study also identified high rates of patient retention and adherence as primary strengths of the clinical program: greater than 99% of patients who were enrolled in the tele-buprenorphine treatment program were retained throughout the duration of their jail stay and were in the program for an average of about two months. Of the 842 patients treated in the program, 27 people were removed for repeated diversion/medication hoarding attempts, representing 3% of the total sample—a relatively small proportion, findings that are compatible with other studies suggesting that diversion does not commonly occur (Evans et al., 2023; Evans, Pivovarova, et al., 2022).

### Limitations and Future Directions

Our study findings should be considered in light of several limitations. Although the multi-site nature of this program is a strength, all jails were in rural regions of a single state; thus, the results and our experiences with providing telemedicine-delivered buprenorphine may not generalize to other areas of the country. Further, the program was implemented in six different jail settings, each of which had their own protocols and approaches to dosing, referrals and other clinical operations. Additionally, data from the jail records were obtained via self-report and were not verified through alternate means. As an observational study of a tele-buprenorphine multi-site jail program, we did not have access to post-release data; thus, the current report focuses only on in-custody care. Future research should focus on linking this in-custody data to external outcomes to build on our previous positive single-site post-release findings (Belcher et al., 2025).

## Data Availability

All data produced in the present study are available upon reasonable request to the authors

## Human Ethics and Consent to Participate declarations

The study was approved as a consent-waivered chart review by the University of Maryland’s (UM) Human Research Protection Office (UMB IRB protocol No. HP - 00100535).

